# Prevalence and clinical correlates of COVID-19 outbreak among health care workers in a tertiary level hospital in Delhi

**DOI:** 10.1101/2020.07.21.20159301

**Authors:** Ankit Khurana, GP Kaushal, Rishi Gupta, Vansh Verma, Kabir Sharma, Manmohan Kohli

## Abstract

In this study, we summarize the epidemiological characteristics of COVID-19 outbreak among Healthcare workers (HCWs) in a tertiary care hospital and compared various parameters and preventive measures taken by positive HCWs to a comparable cohort of COVID negative HCWs. 52.1% of COVID-19 positive HCWs showed symptoms of which only three needed hospitalization possibly due to a younger cohort of HCWs who got infected (35.9 ± 9.3 years). Findings of present study found some protective role of full course prophylactic hydroxychloroquine as compared to a control group (p=0.021) and use of N95 masks over others (p<0.001). Our results did not show any added protection with the use of prophylactic Vitamin C, D, Zinc, or betadine gargles. We also observed outbreak control with increased awareness, near universal testing, PPE provision, sanitization drive, and promoting social distancing among HCWs.

## 1. Introduction

An epidemic caused by Severe Acute Respiratory Syndrome Coronavirus-2 (SARS-CoV-2), a positive sense single stranded RNA virus of zoonotic origin, emerged in Wuhan, Hubei Province, China, in December 2019. This infection has since been spreading rapidly globally, causing significant morbidity and mortality, with COVID-19 cases having been identified in several other countries and territories. WHO has declared it as a public health emergency of international concern. Person-to-person transmission has been widely documented. The transmission of COVID-19 is potent, and the secondary attack rate is high. There is neither a specific drug nor a vaccine for COVID-19. Treatment mainly consists of symptomatic supportive therapy. Over 1,50,000 cases have been detected in India as of 29 May 2020, leading it to become the primary cause of health-related concern in the country.

Governments and health bodies worldwide have been working on pandemic mitigation strategies. These efforts aim to ensure rapid evaluation and care of patients, limiting further transmission, and to better understand risk factors for transmission. With and aim to understand the prevalence and correlates of this infection in a tertiary level hospital in Delhi, we planned the current study. We aimed to look at the infection rate, and the various factors associated with a positive COVID-19 result, which may help in formulating better strategies for preventing the illness.

## 2. Methods

A questionnaire based analysis was carried out to analyze the epidemiological and clinical parameters of healthcare workers of our hospital who had tested positive for COVID-19. A matched cohort of healthcare workers who tested negative was taken as the control group. Data collection was by telephonic surveys, as well as evaluation of health records. Responses were recorded via text, or in-person in certain cases, as the situation allowed. Various epidemiological parameters along with symptoms, co-morbidities, and preventive strategies adopted by healthcare workers were recorded after obtaining due consent. A similar survey was done for the control group.

### 2.1 Statistical analysis

Data was coded and recorded in MS Excel spreadsheet program. SPSS v23 (IBM Corp.) was used for statistical analysis. Normal distribution of data was assessed using the Shapiro-wilk test. Descriptive statistics were elaborated in the form of means for continuous variables, and frequencies and percentages for categorical variables. Group comparisons for continuously distributed data were made using independent sample ‘t’ test when comparing two groups. If data were found to be non-normally distributed, appropriate non-parametric tests were used for these comparisons. Chi-squared test was used for group comparisons for categorical data. In case the expected frequency in the contingency tables was found to be <5 for >25% of the cells, Fisher’s Exact test was used instead. Statistical significance was kept at p < 0.05.

### 2.2 Ethical approval

Data collection and analysis of cases was part of a continuing public health outbreak investigation and was thus considered exempt from institutional review board approval. However, permission was sought from the head of the institute prior to start of the study.

## 3 Results

### 3.1 Demographic data of health care workers

Males constituted 59.6% of total patients in the COVID positive group, whereas 69.0% participants in negative group were male. The mean age in the positive group was 35.98 years while in the negative group was 34.28 years. 16% positive patients were doctors (including faculty and residents), 43.6% were nursing officers, 29.8% were paramedical staff (including sanitary workers, housekeeping staff, and orderlies), and 10.6% security guards. There was no statistically significant difference in terms of age (P=0.231), gender (P=0.188) and designation (P=0.102) between the study and the control groups. (Table 1)

**Table 1:** Depicting demographic data of included healthcare workers

| Parameters | COVID |  | p value |
| --- | --- | --- | --- |
|  | Positive (n = 94) | Negative (n = 87) |  |
| Age | 35.98 ± 9.28 | 34.28 ± 8.97 | 0.231 |
| Gender |  |  | 0.188 |
| Male | 56 (59.6%) | 60 (69.0%) |  |
| Female | 38 (40.4%) | 27 (31.0%) |  |
| Designation |  |  | 0.102 |
| Doctor | 15 (16.0%) | 9 (10.3%) |  |
| Nursing Staff | 41 (43.6%) | 41 (47.1%) |  |
| Paramedical Staff | 28 (29.8%) | 28 (32.2%) |  |
| Security Guard | 10 (10.6%) | 10 (11.5%) |  |

### 3.2 Clinical Presentation

52.1% (49/94) of the participants in the positive group were symptomatic. Fever was the most common symptom, as experienced by 30.9% patients/participants. Other commonly reported symptoms include sore throat, myalgia, headache and cough as reported by 20.2%,20.2%,14.9%,11.7% patients/participants respectively; 4.3% rhinorrhea, 6.4% shortness of breath, 3.2% anosmia, 2.1 % nausea, 2.1% pain abdomen and 1 patient each had mucosal dryness,hemoptysis, dysgeusia and loss of appetite. (Table 2) Three patients in total needed hospitalization and out of the three one patient needed intensive care for recovery. There were no mortality.

**Table 2:** Clinical presentation of COVID positive healthcare workers

| Frequency among COVID positive healthcare workers |  |
| --- | --- |
| Symptomatic |  |
| Yes | 49 (52.1%) |
| No | 45 (47.9%) |
| Maybe | 0 (0.0%) |
| Fever | 29 (30.9%) |
| Cough | 11 (11.7%) |
| Sore throat | 19 (20.2%) |
| Runny nose/Rhinorrhea | 4 (4.3%) |
| Mucosal Dryness | 1 (1.1%) |
| Hemoptysis | 1 (1.1%) |
| Breathlessness/ pneumonia/ chest congestion | 6 (6.4%) |
| Chest pain | 0 (0.0%) |
| Bodyache/Myalgia/ arthralgia/fatigue/weakness/malaise | 19 (20.2%) |
| Pain abdomen | 2 (2.1%) |
| Headache | 14 (14.9%) |
| Dysphagia | 1 (1.1%) |
| Diarrhea | 0 (0.0%) |
| Anosmia | 3 (3.2%) |
| Loss of appetite | 1 (1.1%) |
| Loss of taste/alterd taste | 1 (1.1%) |
| Nausea/vomiting | 2 (2.1%) |

### 3.3 Contact analysis

The mean number of close contacts per workday (>6 hours) in the COVID Positive group was 19.13 (21.50) and in the Negative group was 17.85 (13.70). The close contacts per workday in the COVID Positive ranged from 0-150 . The close contacts per workday in the COVID: Negative/Unknown ranged from 0-60. There was no significant difference between the groups in terms of close contacts per workday (W = 3794.500, p = 0.402). None of the positive participants had suspected contact at home. (Table 3)

**Table 3.** Contact analysis for COVID positive and control group

|  | COVID Positive | COVID Positive | P value |
| --- | --- | --- | --- |
| Close Contacts per Workday (>6 hours) | 19.13 ± 21.50 | 17.85 ± 13.70 | 0.402 |
| Place of Suspected Contact |  |  |  |
| Home | 0 (0.0%) | - |  |
| Hospital | 94 (100.0%) | - |  |
| Duration of Contact |  |  |  |
| <6 hours | 32 (51.6%) | - |  |
| >6 hours | 30 (48.4%) | - |  |

### 3.4 Co-morbidity analysis

10.6% (14/94) of the participants in the positive group suffered from chronic medical condition, while 12.6% (14/87) of the participants in the control group had chronic medical conditions. The difference between the two groups was not statistically significant (p=0.955) using non-parametric tests (Wilcoxon-Mann-Whitney U Test). The Total number of co-morbidities in the positive ranged from 0 – 3 while in the negative group from 0–4. (Table 4)

**Table 4.**
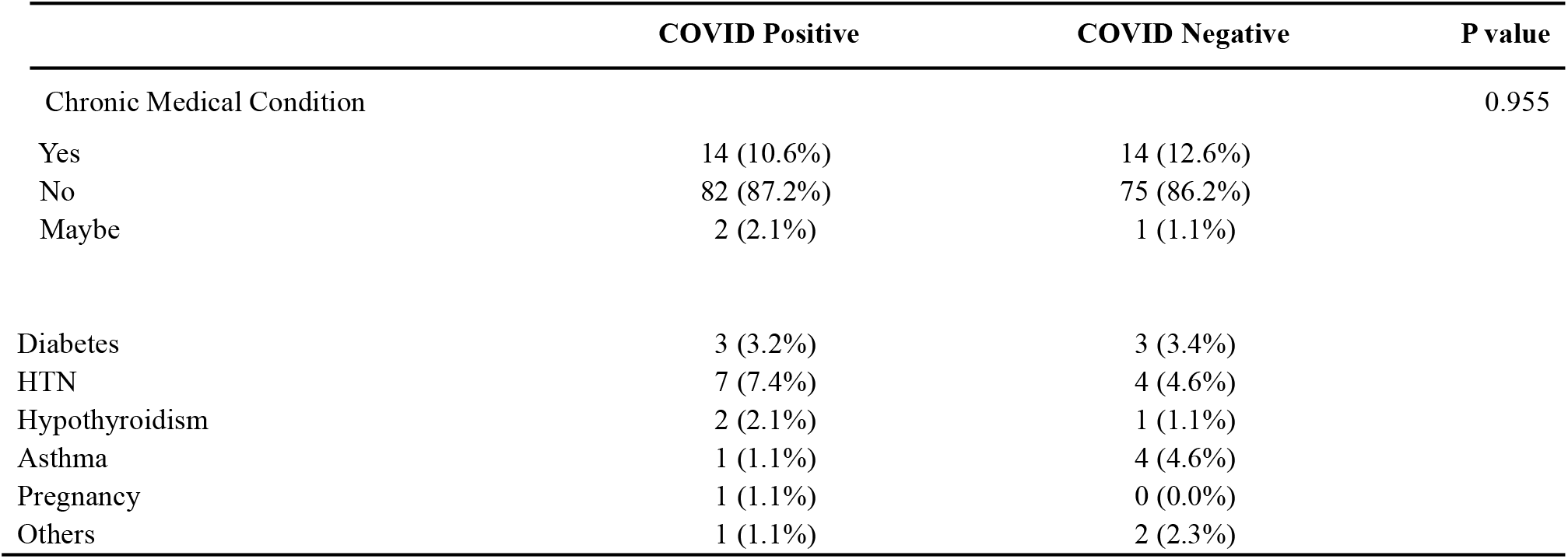
Distribution of chronic medical conditions among positive healthcare workers and control group

|  | COVID Positive | COVID Negative | P value |
| --- | --- | --- | --- |
| Chronic Medical Condition |  |  | 0.955 |
| Yes | 14 (10.6%) | 14 (12.6%) |  |
| No | 82 (87.2%) | 75 (86.2%) |  |
| Maybe | 2 (2.1%) | 1 (1.1%) |  |
| Diabetes | 3 (3.2%) | 3 (3.4%) |  |
| HTN | 7 (7.4%) | 4 (4.6%) |  |
| Hypothyroidism | 2 (2.1%) | 1 (1.1%) |  |
| Asthma | 1 (1.1%) | 4 (4.6%) |  |
| Pregnancy | 1 (1.1%) | 0 (0.0%) |  |
| Others | 1 (1.1%) | 2 (2.3%) |  |

### 3.5 Prophylactic agents used by healthcare workers

Chi-squared test was used to determine the association between ‘COVID positivity’ and ‘prophylactic hydroxychloroquine Intake’. There was a significant difference between the various groups in terms of distribution of prophylactic hydroxychloroquine Intake (*X*^*2*^ = 17.159, p =<0.001). 6.4% of the participants in the positive group had taken full course of hydroxychloroquine of 7 weeks or more. 18.4% of the participants in the negative group had taken the full course of hydroxychloroquine prophylaxis. An analysis between those who had taken full course and those who had taken either incomplete course or had not taken at all revealed a statistically significant difference with p = 0.021 using the Fisher’s exact test. (Table 5 and 6)

**Table 5:**
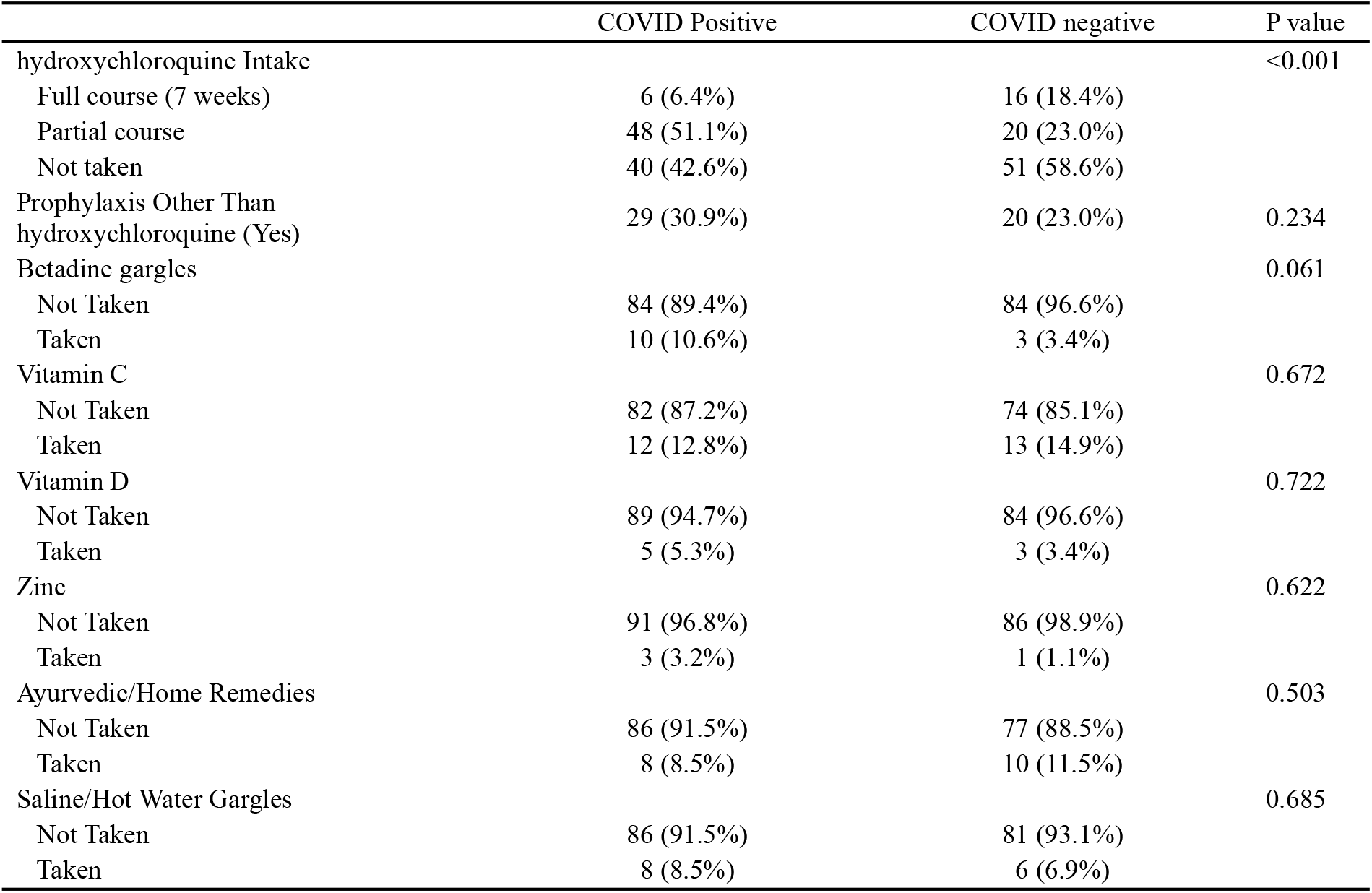
Analysis of preventive and prophylactic strategies adopted by healthcare workers.

|  | COVID Positive | COVID negative | P value |
| --- | --- | --- | --- |
| hydroxychloroquine Intake |  |  | <0.001 |
| Full course (7 weeks) | 6 (6.4%) | 16 (18.4%) |  |
| Partial course | 48 (51.1%) | 20 (23.0%) |  |
| Not taken | 40 (42.6%) | 51 (58.6%) |  |
| Prophylaxis Other Than hydroxychloroquine (Yes) | 29 (30.9%) | 20 (23.0%) | 0.234 |
| Betadine gargles |  |  | 0.061 |
| Not Taken | 84 (89.4%) | 84 (96.6%) |  |
| Taken | 10 (10.6%) | 3 (3.4%) |  |
| Vitamin C |  |  | 0.672 |
| Not Taken | 82 (87.2%) | 74 (85.1%) |  |
| Taken | 12 (12.8%) | 13 (14.9%) |  |
| Vitamin D |  |  | 0.722 |
| Not Taken | 89 (94.7%) | 84 (96.6%) |  |
| Taken | 5 (5.3%) | 3 (3.4%) |  |
| Zinc |  |  | 0.622 |
| Not Taken | 91 (96.8%) | 86 (98.9%) |  |
| Taken | 3 (3.2%) | 1 (1.1%) |  |
| Ayurvedic/Home Remedies |  |  | 0.503 |
| Not Taken | 86 (91.5%) | 77 (88.5%) |  |
| Taken | 8 (8.5%) | 10 (11.5%) |  |
| Saline/Hot Water Gargles |  |  | 0.685 |
| Not Taken | 86 (91.5%) | 81 (93.1%) |  |
| Taken | 8 (8.5%) | 6 (6.9%) |  |

**Table 6:** Distribution of hydroxychloroquine prophylaxis usage among healthcare workers

| Hydroxychloroquine | COVID status |  | P Value |
| --- | --- | --- | --- |
|  | Negative | Positive |  |
| Full course | 16 | 6 | 0.012 |
| Not taken/Incomplete course | 71 | 88 |  |

Commonly used prophylactic agents other than hydroxychloroquine included Betadine gargles, Vitamin C, D, Zinc, saline gargles, and other home remedies did not reveal significant association with COVID status among healthcare workers. (p>0.05). 30.9% of the participants in the positive group used prophylaxis other than hydroxychloroquine while in the negative group 23.0% of the participants used other prophylaxis. (Table 5)

95.7% of the participants in the COVID positive healthcare workers used mask at both work and community. Similarly, 90.8% of the participants in the control group had mask usage at both places. Chi-squared test was used to explore the association between ‘COVID status’ and place of mask usage. We couldn’t find a significant difference (*X*^*2*^ = 1.781, p = 0.182). Chi-squared test was further used to explore the association between ‘COVID’ and ‘Minimum level of protection’.

A significant difference was seen in terms of minimum level of protection (*X*^*2*^= 15.668, p = <0.001). 30.9% of the participants in the positive group had minimum level of protection as an N95 mask. 63.8% of the participants in the positive group had minimum level of protection as a 3-Ply Mask, whereas 5.3% of the participants in the group positive group had minimum level of protection as a bandana. The percentages in the control group were 57.5%, 34.5% and 8.0% for N95, 3ply and Bandana mask, respectively. Thus, control group had the larger fraction of people using N95 as compared to 3 ply and bandana (p<0.001) thus a significantly higher number of participants were using M-95 masks as compared to bandana in the control group.

Average days after which N95 mask was changed was not normally distributed in the 2 groups. Thus, non-parametric tests (Wilcoxon-Mann-Whitney U Test) were used to compare groups. The mean (SD) number of days before mask was changed in the COVID Positive group was 7.81 (10.53) while in the negative group was 7.33 (9.23). No significant difference between the groups in terms of N95 mask change frequency (W = 3821.500, p = 0.439) was observed. (Table 7 and 8)

**Table 7:** Analysis of mask usage among healthcare workers.

|  | COVID positive | COVID negative | P value |
| --- | --- | --- | --- |
| Mask Use at Work only) | 3 (3.2%) | 7 (8.0%) | 0.199 |
| Mask Use in Community only | 1 (1.1%) | 1 (1.1%) | 1.000 |
| Mask Use at Both Places | 90 (95.7%) | 79 (90.8%) | 0.182 |
| N95 Mask Use (Present) | 62 (66.0%) | 77 (88.5%) | <0.001 |
| 3-Ply Mask Use (Present) | 62 (66.0%) | 34 (39.1%) | <0.001 |
| Bandana Use (Present) | 5 (5.3%) | 3 (3.4%) | 0.722 |
| Minimum level of protection |  |  | <0.001 |
| N95 | 29 (30.9%) | 50 (57.5%) |  |
| 3-Ply Mask | 60 (63.8%) | 30 (34.5%) |  |
| Bandana | 5 (5.3%) | 7 (8.0%) |  |
| Mask change frequency(no. of days each mask used) | 7.81 ± 10.53 | 7.33 ± 9.23 | 0.439 |
| Mask Use at Time of Contact |  |  | <0.001 |
| Yes | 58 (61.7%) |  |  |
| No | 0 (0.0%) |  |  |
| Maybe | 36 (38.3%) |  |  |

Significant at $p < 0.05$

**Table 8:**
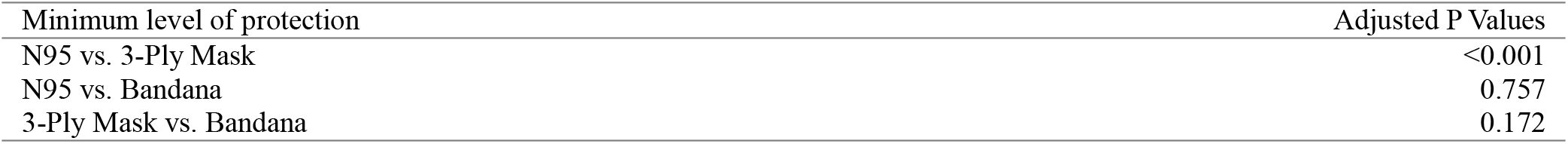
Analysis of mask used as minimum level of protection among healthcare workers.

| Minimum level of protection | Adjusted P Values |
| --- | --- |
| N95 vs. 3-Ply Mask | <0.001 |
| N95 vs. Bandana | 0.757 |
| 3-Ply Mask vs. Bandana | 0.172 |

### 3.7 Hand Hygiene practices among Health care Workers

Participants in the group COVID positive group had the larger proportion of people who washed hands for over 20 seconds than the control group, but the difference did not attain significance. The variable Handwashing/Sanitizer-Use Frequency (Times/day) was not normally distributed in the two groups. Thus, non-parametric tests (Wilcoxon-Mann-Whitney U Test) were used to make group comparisons. The mean (SD) of Handwashing/Sanitizer-Use Frequency (Times/day) in the positive group was 20.66 (12.79) and in control group was 18.03 (13.16). There was no significant difference between the groups in terms of Handwashing/Sanitizer-Use Frequency (Times/day) (W = 4633.000, p = 0.120). (Table 9)

**Table 9.** Analysis of hand hygiene practices among health care workers

|  | COVID Positive | COVID Negative | P value |
| --- | --- | --- | --- |
| Hand washing Technique |  |  | 0.243 |
| Normal (<20s) | 30 (31.9%) | 52 (59.8%) |  |
| Surgical (>20s) | 64 (68.1%) | 35 (40.2%) |  |
| Handwashing/Sanitizer-Use Frequency (Times/day) | 20.66 ± 12.79 | 18.03 ± 13.16 | 0.120 |

## 4. Discussion

We have provided an initial assessment of the epidemiology and risk mitigation dynamics of the COVID-19 outbreak among healthcare workers of our hospital. We observed a marked increase in positive cases within a few days, of which only three needed hospitalization and one required admission to intensive care unit. Only 52.1% of the positive healthcare workers showed symptoms. A possible reason for the same is believed to be younger cohort of patients amongst the healthcare workers who got infected. Moreover, we found that the majority (about 60%) of COVID-19 cases were male, although the reason remains to be clarified.

Findings of present study found some protective role of full course (7 weeks) of prophylactic hydroxychloroquine as compared to a control group of negative healthcare workers with p=0.021 and use of N95 masks over others. Our results did not show any added protection with the use of other strategies in the form of prophylactic Vitamin C, D, Zinc, betadine gargles, or any other home remedy.

### 4.1 Infectivity and Transmission

Estimation of the prevalence and transmission for undocumented novel coronavirus (SARS-CoV-2) infections is critical for understanding the total prevalence and pandemic potential. Another factor is the mode of viral transmission through aerosol and fomites. Van Doremalen found that SARS-CoV-2 was viable for over 3 hours in aerosol mode but titers decreased significantly after 3 hours. Virus is much stable on stainless steel and plastic compared to copper and cardboard and can be obtained from steel and plastic surfaces even after 3 days.(1) However, till date there is no consensus on Airborne transmission as contradictory versions exist. The matter remains unresolved partially due to the difference in definitions of air borne transmission, different conditions used by various researchers and infectivity if Viral RNA particles in air.(2) Lauer took data of 181 patients and estimated that the median incubation period as 5.1 days which is similar to SARS and 2.5 times that of influenza virus. They found that less than 2.5 % of cases became symptomatic within 2.2 days of exposure and 97.5 % became symptomatic within 11.5 days of exposure. 1/100 developed symptoms after 14 days of exposure.(3)

SARS-CoV-2 shows high infectivity in the hospital setting. It has been isolated from sputum, nasopharynx, oropharynx, stool, blood, & conjunctiva but not from urine, breastmilk, amniotic fluid and cord blood.(4–8) The highest load was found at onset of symptoms and 30% persisted to have RNA load detected by RT-PCR after 20 days of symptom onset.(8) Rapid decline in viral load follows seroconversion which occurs at 6-12 days.(9) Furthermore, Liu et al found that severe cases have higher viral loads and shed the virus longer explaining the high infectivity in hospital settings. Viral load is an indicator of disease severity.(10) Cereda found the pre-symptomatic individuals had viral loads comparable to symptomatic individuals with no significant statistical difference.(11) Wang JID identified 55 family members of known COVID patients and found that RT-PCR positivity 1-7 days before symptom onset. Most patients who were likely to have mild symptoms were younger which is similar to our series where the mean age is 35 years and only over 50% displayed any symptoms.(12) Hu found that truly “asymptomatic” individuals had a mean age of 14 years.(13) This is in contrast to the diamond princess cruise ship cohort where the asymptomatic population which was RT-PCR positive was 17.9%. However, patients were not followed long enough to assess how many of them were pre-symptomatic.(14) Asymptomatic transmission was reported early during the course of the epidemic by Bai and Qian.(15,16) This could be a reason for rapid spread of the virus among healthcare workers, where a patient or co-workers can be asymptomatic carriers for COVID.

Du et al published evaluated 462 transmission events and serial interval (time between symptom onset between transmitter and transmitee) and found it to be 4 days and they reported 6% of transmissions are pre-symptomatic.(17) Epidemic control becomes challenging due to undocumented infections. In China, as high as 86% of infections were undocumented during early epidemic which were either mild or asymptomatic. Undocumented infections were estimated to be half as infectious as symptomatic infections.(18) Environmental viability was assessed and 13 patients with mild COVID 19 were isolated in negative pressure rooms. The authors found viral RNA on cell phones with 85% positivity as well as toilets and air samples. 63 % of air samples were positive for RNA but the live virus could not be cultured. Hence hinting at regular aerosolization of the virus.(1) The possibility of airborne transmission adds complexity to infection control strategies especially in the setting of tertiary hospitals with a large number of positive patients.

Peak infectiousness occurs about 1 day prior to symptom onset. When viremia is assessed Viral loads as previously discussed the viral load is high even in the pre-symptomatic phase and peaks at symptom onset. (19) He at al studied viral shedding by looking at 414 swabs from 94 patients of positive patients from symptom onset. None of them had severe symptoms on presentation although 18 developed the same during the course. They found viral load does not correlate with sex, severity, and age. They also evaluated 77 detailed transmission pairs based on serial interval (that is time from symptom onset in transmitter to symptom onset in transmittee) and they estimated that infectiousness started at 2.3 days prior to symptom onset and peaked 0.7 days prior to symptom onset. Estimated pre-symptomatic transmission was 44% in their study.(20) This is in line with the findings of Du et al and Ferretti et al.(17,21) The authors of this study also suggest that this percentage will vary depending on the efficacy of active case finding with those places with a high percentage of case finding to have a higher percentage of presymptomatic transmission. Huang and Lin et al found that higher viral loads are found in lower respiratory specimens in critically ill.(22) Xiao et al have found in their study of 56 patients that prolonged shedding of viral RNA can be seen in mild or moderate disease and found that Nasopharyngeal PCR remains positive a median of 24 days after symptom onset with a 5% positivity at 5 weeks, however its relation with infectiousness remains unknown(23). In mild cases, live virus isolated up to day 8 after symptom onset. In a study from Taiwan, the authors found that no secondary cases were identified from exposures occurring after 5 days from symptom onset.(24) RNA detection by PCR maybe for much longer.(24) For critical illnesses the scenario can be slightly different and there can be prolonged shedding of viral RNA lasting many weeks, hence leading to high infectivity from critically ill patients in the hospital. (25)

### 4.2 Clinical features

There is a spectrum of presentations for COVID-19 which include: asymptomatic, mildly symptomatic, severe symptomatic with spontaneous recovery, and severe symptomatic with development of an ARDS - proinflammatory syndrome.(23,26,27) Compiled series of hospitalized patients revealed fever and cough to be the most common symptoms followed by myalgias, fatigue, sore throat, nausea, vomiting, diarrhea, headache, and rhinorrhea (22,28)(12,27–29) Gastrointestinal (GI) symptoms have been given more emphasis in recent data. Pan et al found that 42% had GI symptoms as part of their syndrome.(29) Anosmia, hyposmia, and dysgeusia have been reported as per society reports that state that anecdotal evidence is accumulating. A large-scale real-time symptom monitoring study seems to confirm the importance of anosmia/ageusia. (30) Our findings are in line with available literature with most common symptom being fever, followed by bodyache, sore throat, cough, headache, pain abdomen, anosmia and in more severe cases-breathlessness and need for mask ventilation.

### 4.3 Comorbidity evaluation

Risk factors for severe disease as per several authors are comorbidities like hypertension, heart disease, diabetes, CKD, Pre-existing pulmonary disease. Case fatality rate is defined as number of COVID 19 deaths divided by the total number of infections. It is subject to bias if the number of cases tested is too low (upward bias) or when deaths have not yet occurred in setting of ongoing illness (downward bias). Wu et al used this information to assess age specific CFR as 1.4% with 0.3% under 30 and 2.6% over 59 and increasing precipitously as age increases.(31) The present study did not find any severe illness and only 3/94 positive patients needed hospital admission. In contrast, mean age of infected health care workers in our study was 35 years and only 14/94 (14.9%) had one or more co-morbidities.

### 4.4 Hydroxychloroquine prophylaxis

The Indian Council of Medical Research, under the Ministry of Health and Family Welfare, has recommended chemoprophylaxis with hydroxychloroquine (400 mg twice on day 1, then 400 mg once a week thereafter for 7 weeks) for asymptomatic healthcare workers treating patients with suspected or confirmed COVID-19, and for asymptomatic household contacts of confirmed cases. (32)Our results show a significant reduction in the rate of infection by taking full course of hydroxychloroquine prophylaxis, however accumulating therapeutic data shows no benefit. Hydroxychloroquine was found to have in vitro activity early on in the pandemic. It binds to ACE2 receptor and as a result may block viral entry and intracellular transfer.(33) An interim report with a rather uncertain methodology from china reported efficacy and safety in 100 patients leading to expert consensus in china to recommend chloroquine as the first line for all patients with COVID 19. Low quality RCTs from china showed no significant effect of HCQ with respect to negative throat swab, days to being afebrile and radiographic progression. (34,35) The bias was that both groups received other anti-virals that is lopinavir and ritonavir. Chen et al in another RCT suggested superior clinical results in hydroxychloroquine treated individuals but the study has several biases in terms of 80 excluded patients for uncertain reasons and it is unclear how other standard therapies were distributed among hydroxychloroquine and non-hydroxychloroquine groups.(36) A small non-randomized french clinical trial (26 HCQ vs 16 HCQ) suggested faster viral clearance with hydroxychloroquine, but they excluded 6 patients from the analysis who were either shifted to ICU or got worse which suggests confounding in their data.(37,38) The society (International society of antimicrobial chemotherapy) that published this paper said in a statement that the published study did not meet the societies expected standard relating to a lack of better explanation of inclusion criteria.

The same group of French authors (Gautret et al) have published synergy between azithromycin and hydroxychloroquine based on faster viral clearance when a combination of the two drugs was used.(37) The limitation of their findings were - no control arm and only patients with mild disease were included. (38) Mahevas did a retrospective analysis of 181 patients from 4 hospitals of which 84 who had taken hydroxychloroquine and 79 did not. The authors found no benefit and approximately 10% needed to discontinue hydroxychloroquine due to QT_C_ prolongation issue. (39) Tang conducted a multi-centric RCT of 150 hospitalized patients with COVID and found that hydroxychloroquine doesn’t lead to faster viral clearance. Post hoc analysis controlling for use of other antivirals found early symptom relief with hydroxychloroquine and faster improvement in CRP. (40) Magagnoli retrospectively analyzed 368 males. They found that in their non-randomized study found that risk of death was higher with hydroxychloroquine group. (41)

### 4.5 Mask usage

A recent study on influenza referenced above shows no difference lab confirmed influenza with use of either N95 or surgical mask, influenza virus like COVID is an RNA virus with similar infectivity.(42) Leung assessed the use of masks in transmission of respiratory viruses including Influenza, non COVID Coronaviruses, and Rhinoviruses between 2013 to 2016. They included 246 participants with respiratory symptoms who had confirmed respiratory virus and randomized 50% to wear masks and assessed exhaled breath particles and classified them as either droplets or aerosols. There was a significant decrease in recovery of coronaviruses from droplets and aerosols with the use of masks. While this study was not done in context of COVID 19 but the findings are extrapolatable to COVID patients especially those who are symptomatic. (43)

We, however, found N95 to be more effective in preventing infection as compared to other masks (3 ply mask and cloth bandana). (p<0.001) There was no difference in frequency of N95 mask change among positive and control groups and most healthcare workers who tested positive did show good compliance with mask usage in the hospital as well as in the community.

### 4.6 Outbreak control

#### 4.6.1 Situation appropriate personal protective equipment

McMichael et al explained transmission and outcome s in an epidemic at an unprepared long-term care facility. 81/130 residents, 34/170 health care workers and 14 visitors became positive. The median age of 81 residents was high with most having several co-morbidities and eventually 27 % died. The contributing factors analyzed by the authors were: Staff members worked while symptomatic, Staff members worked in more than one facility, Inadequate familiarity and adherence to standard, droplet, and contact precautions and eye protection recommendations, challenges to implementing infection control practices including inadequate supplies of PPE and other items (e.g. Hand sanitizer), delayed recognition of cases because of low index of suspicion, limited testing availability, and difficulty identifying persons with COVID-19 based on signs and symptoms alone. The authors concluded that “Long-term care facilities should take proactive steps to protect the health of residents and preserve the health care workforce by identifying and excluding potentially infected staff members and visitors, ensuring early recognition of potentially infected patients, and implementing appropriate infection control measures”.(44,45) The hospital administration taking cognizance of above research took pro-active steps to contain the outbreak among health care workers. All health care workers who tested positive were immediately instructed to isolate themselves. Similarly, high risk contacts of positive individuals were sent on quarantine pro-actively and timely. Situation appropriate PPEs were provided, and all health care workers were instructed to adhere to given rules for their protection. A surveillance was carried out and most of the 1605 health care workers employed at our facility were tested. This led to further case identification and lead to contact tracing, quarantine of close contacts and control of the outbreak.

#### 4.6.2 Movement restriction

Movement restrictions and mandatory social distancing are highly effective in controlling transmission and decrease transmission coefficient from 4 to 1.6 (and later less than 1 when quarantine is initiated).(45)The institute administration promoted mandatory social distancing and necessary quarantine/isolation in our mitigation strategy. This led to control of the initial outbreak among our health care workers. Lower case reporting was achieved subsequently.

#### 4.6.3 Contact tracing

Ferretti et al studied the impact of 2 interventions which were contact tracing and case identification. Their model explains that if there is a delay in isolation and contact quarantine, there is less likelihood of it being effective at controlling the epidemic. The authors also support the use of a mobile phone app which is a form of digital contact tracing by keeping a temporary record of proximity events between individuals. This approach would not require coercive surveillance since the system can achieve epidemic suppression. This is a promising strategy. Epidemic control becomes feasible with contact tracing if minimal delay can be achieved.(21) The hospital administration’s approach in the present scenario was comparable. Rapid contact tracing was initiated, and all contacts and positive individuals were encouraged to download government of India’s contact tracing app (Arogya Setu) immediately when the outbreak was identified.

#### 4.6.4 Universal Testing drive

Gudbjartsson found that targeted sampling was changed to random sampling in Ireland which was an open invitation to asymptomatic or mild cold symptoms. The authors found <1% had positive SARS-CoV-2 PCR.(46) Lavezzo assessed role of mass testing in a town in Italy with a population of 3000. The entire town was quarantined for 14 days after the index case had died. The researchers found high rate of asymptomatic infection and transmission upon universal testing. Two PCR samples were taken 12 days apart and the authors found that 85% and 71% population took part in the survey. Prevalence decreased to 1.2% the second time from an initial 2.6%t. A quarter of those positive on the second time point were new infections which the researchers found were due to contact with asymptomatic infected individuals either before lockdown or within their household. This substantiates the point that asymptomatic/pre-symptomatic transmission plays a key role in ongoing transmission of SARS-CoV-2. The effective reproductive number declined from 3 to 0.14 after the end of lockdown - effectively ending the outbreak. (47) Studies with near -universal screening of various populations are now becoming more available, finding a wide range of asymptomatic people with positive PCR Pregnant women in NYC-13.5% (87% of total infections); homeless shelter in Boston - 36% (great majority of total infections); Town in Italy - <1% (41% of total infections); Iceland - <1% (43% of total infections) and cruise ship Diamond Princess - 9% (46% of total infections) (48–50) Thus, serological data shows SARS-CoV-2 has a significant iceberg effect. Our approach to control involved near universal testing of health care workers to understand the level of the outbreak and early outbreak control.

#### 4.6.5 Rigorous sanitization drive

Liu measured viral RNA in various areas in 2 hospitals. They found that ventilation and sanitization played a significant role in the number of viral RNA copies detected. Ventilated areas had a lower RNA concentration as compared to unventilated areas like toilets. Also, a rigorous sanitization drive lead to virtually undetectable detection of viral RNA. (51) This lead the hospital authorities to undertake a massive sanitization drive in the hospital.

## 5. Conclusion

The transmission potential of COVID-19 is very high, and the number of cases may become largely unsustainable for the healthcare system in a very short-time horizon. It is important to protect the healthcare force for effective epidemic management. We observed outbreak control with increased awareness, near universal testing, PPE provision, sanitization drive, and promoting social distancing among health care workers. This study also brings forth the role of hydroxychloroquine prophylaxis, use of N95 mask, and effect of early interventions in outbreak mitigation. Aggressive containment strategies are required to control COVID-19 spread and catastrophic outcomes for the healthcare system in the absence of a therapy/vaccine to avoid overwhelming the critical care capacity of any healthcare facility.

## Data Availability

data associated with a paper is available, and data can be accessed after writing to the corresponding author

## Notes

### Competing Interest Statement

The authors have declared no competing interest.

### Funding Statement

No funding was received or used

### Author Declarations

Institute ethical and scientific committee, BSAMCH

## References

1. van Doremalen N, Bushmaker T, Morris DH, Holbrook MG, Gamble A, Williamson BN, et al. Aerosol and Surface Stability of SARS-CoV-2 as Compared with SARS-CoV-1. N Engl J Med. 2020 Apr 16;382(16):1564–7.

2. Ong SWX, Tan YK, Chia PY, Lee TH, Ng OT, Wong MSY, et al. Air, Surface Environmental, and Personal Protective Equipment Contamination by Severe Acute Respiratory Syndrome Coronavirus 2 (SARS-CoV-2) From a Symptomatic Patient. JAMA. 2020 Apr 28;323(16):1610.

3. Lauer SA, Grantz KH, Bi Q, Jones FK, Zheng Q, Meredith HR, et al. The Incubation Period of Coronavirus Disease 2019 (COVID-19) From Publicly Reported Confirmed Cases: Estimation and Application. Annals of Internal Medicine. 2020 May 5;172(9):577–82.

4. Zou L, Ruan F, Huang M, Liang L, Huang H, Hong Z, et al. SARS-CoV-2 Viral Load in Upper Respiratory Specimens of Infected Patients. N Engl J Med. 2020 Mar 19;382(12):1177–9.

5. Pan Y, Zhang D, Yang P, Poon LLM, Wang Q. Viral load of SARS-CoV-2 in clinical samples. Lancet Infect Dis. 2020;20(4):411–2.

6. Wang W, Xu Y, Gao R, Lu R, Han K, Wu G, et al. Detection of SARS-CoV-2 in Different Types of Clinical Specimens. JAMA. 2020 Mar 11;

7. Liang L, Wu P. There may be virus in conjunctival secretion of patients with COVID-19. Acta Ophthalmol. 2020 May;98(3):223.

8. To KK-W, Tsang OT-Y, Leung W-S, Tam AR, Wu T-C, Lung DC, et al. Temporal profiles of viral load in posterior oropharyngeal saliva samples and serum antibody responses during infection by SARS-CoV-2: an observational cohort study. Lancet Infect Dis. 2020;20(5):565–74.

9. Woelfel R, Corman VM, Guggemos W, Seilmaier M, Zange S, Mueller MA, et al. Clinical presentation and virological assessment of hospitalized cases of coronavirus disease 2019 in a travel-associated transmission cluster [Internet]. Infectious Diseases (except HIV/AIDS); 2020 Mar [cited 2020 May 27]. Available from: http://medrxiv.org/lookup/doi/10.1101/2020.03.05.20030502

10. Liu Y, Yan L-M, Wan L, Xiang T-X, Le A, Liu J-M, et al. Viral dynamics in mild and severe cases of COVID-19. Lancet Infect Dis. 2020 Mar 19;

11. D C, M T, F R, V D, M A, P P, et al. The early phase of the COVID-19 outbreak in Lombardy, Italy. arXiv:200309320 [q-bio] [Internet]. 2020 Mar 20 [cited 2020 Jun 5]; Available from: http://arxiv.org/abs/2003.09320

12. Wang Y, Liu Y, Liu L, Wang X, Luo N, Li L. Clinical Outcomes in 55 Patients With Severe Acute Respiratory Syndrome Coronavirus 2 Who Were Asymptomatic at Hospital Admission in Shenzhen, China. The Journal of Infectious Diseases. 2020 May 11;221(11):1770–4.

13. Hu Z, Song C, Xu C, Jin G, Chen Y, Xu X, et al. Clinical characteristics of 24 asymptomatic infections with COVID-19 screened among close contacts in Nanjing, China. Sci China Life Sci. 2020;63(5):706–11.

14. Mizumoto K, Kagaya K, Zarebski A, Chowell G. Estimating the asymptomatic proportion of coronavirus disease 2019 (COVID-19) cases on board the Diamond Princess cruise ship, Yokohama, Japan, 2020. Euro Surveill. 2020;25(10).

15. Bai Y, Yao L, Wei T, Tian F, Jin D-Y, Chen L, et al. Presumed Asymptomatic Carrier Transmission of COVID-19. JAMA. 2020 Feb 21;

16. Qian G, Yang N, Ma AHY, Wang L, Li G, Chen X, et al. A COVID-19 Transmission within a family cluster by presymptomatic infectors in China. Clin Infect Dis. 2020 Mar 23;

17. Du Z, Xu X, Wu Y, Wang L, Cowling BJ, Meyers LA. Serial Interval of COVID-19 among Publicly Reported Confirmed Cases. Emerging Infect Dis. 2020;26(6):1341–3.

18. Li R, Pei S, Chen B, Song Y, Zhang T, Yang W, et al. Substantial undocumented infection facilitates the rapid dissemination of novel coronavirus (SARS-CoV-2). Science. 2020 01;368(6490):489–93.

19. Zhou F, Yu T, Du R, Fan G, Liu Y, Liu Z, et al. Clinical course and risk factors for mortality of adult inpatients with COVID-19 in Wuhan, China: a retrospective cohort study. Lancet. 2020 28;395(10229):1054–62.

20. He X, Lau EHY, Wu P, Deng X, Wang J, Hao X, et al. Temporal dynamics in viral shedding and transmissibility of COVID-19. Nat Med. 2020 May;26(5):672–5.

21. Ferretti L, Wymant C, Kendall M, Zhao L, Nurtay A, Abeler-Dörner L, et al. Quantifying SARS-CoV-2 transmission suggests epidemic control with digital contact tracing. Science. 2020 08;368(6491).

22. Huang C, Wang Y, Li X, Ren L, Zhao J, Hu Y, et al. Clinical features of patients infected with 2019 novel coronavirus in Wuhan, China. Lancet. 2020 15;395(10223):497–506.

23. Xiao AT, Tong YX, Zhang S. Profile of RT-PCR for SARS-CoV-2: a preliminary study from 56 COVID-19 patients. Clin Infect Dis. 2020 Apr 19;

24. Cheng H-Y, Jian S-W, Liu D-P, Ng T-C, Huang W-T, Lin H-H, et al. Contact Tracing Assessment of COVID-19 Transmission Dynamics in Taiwan and Risk at Different Exposure Periods Before and After Symptom Onset. JAMA Intern Med [Internet]. 2020 May 1 [cited 2020 May 28]; Available from: https://jamanetwork.com/journals/jamainternalmedicine/fullarticle/2765641

25. Arons MM, Hatfield KM, Reddy SC, Kimball A, James A, Jacobs JR, et al. Presymptomatic SARS-CoV-2 Infections and Transmission in a Skilled Nursing Facility. N Engl J Med. 2020 May 28;382(22):2081–90.

26. Siddiqi HK, Mehra MR. COVID-19 illness in native and immunosuppressed states: A clinical-therapeutic staging proposal. J Heart Lung Transplant. 2020;39(5):405–7.

27. Pan X, Chen D, Xia Y, Wu X, Li T, Ou X, et al. Asymptomatic cases in a family cluster with SARS-CoV-2 infection. Lancet Infect Dis. 2020;20(4):410–1.

28. Guan W, Ni Z, Hu Y, Liang W, Ou C, He J, et al. Clinical Characteristics of Coronavirus Disease 2019 in China. N Engl J Med. 2020 Apr 30;382(18):1708–20.

29. Pan L, Mu M, Yang P, Sun Y, Wang R, Yan J, et al. Clinical Characteristics of COVID-19 Patients With Digestive Symptoms in Hubei, China: A Descriptive, Cross-Sectional, Multicenter Study. Am J Gastroenterol. 2020;115(5):766–73.

30. Reinhard A, Ikonomidis C, Broome M, Gorostidi F. [Anosmia and COVID-19]. Rev Med Suisse. 2020 Apr 29;16(N° 691-2):849–51.

31. Wu X, Nethery RC, Sabath BM, Braun D, Dominici F. Exposure to air pollution and COVID-19 mortality in the United States: A nationwide cross-sectional study [Internet]. Epidemiology; 2020 Apr [cited 2020 May 28]. Available from: http://medrxiv.org/lookup/doi/10.1101/2020.04.05.20054502

32. ICMR. National Taskforce for COVID-19. Advisory on the use of hydroxy-chloroquine as prophylaxis for SARS-CoV-2 infection. 2020. [Internet]. Available from: https://www.mohfw.gov.in/pdf/AdvisoryontheuseofHydroxychloroquinasprophylaxisforSARSCoV2infection.pdf

33. Chen Jun, LIU Danping,Liu Li, Liu Ping, Xu Qingnian, Xia Lu, Ling Yun, Huang Dan, SONG Shuli, Zhang Dandan, Qian Zhiping, Li Tao, Shen Yinzhong, Lu Hongzhou. A pilot study of hydroxychloroquine in treatment of patients with moderate COVID-19. J Zhejiang Univ (Med Sci). 2020;49(2):215–9.

34. Gao J, Tian Z, Yang X. Breakthrough: Chloroquine phosphate has shown apparent efficacy in treatment of COVID-19 associated pneumonia in clinical studies. Biosci Trends. 2020 Mar 16;14(1):72–3.

35. Wang M, Cao R, Zhang L, Yang X, Liu J, Xu M, et al. Remdesivir and chloroquine effectively inhibit the recently emerged novel coronavirus (2019-nCoV) in vitro. Cell Res. 2020;30(3):269–71.

36. Chen Z, Hu J, Zhang Z, Jiang S, Han S, Yan D, et al. Efficacy of hydroxychloroquine in patients with COVID-19: results of a randomized clinical trial [Internet]. Epidemiology; 2020 Mar [cited 2020 May 28]. Available from: http://medrxiv.org/lookup/doi/10.1101/2020.03.22.20040758

37. Gautret P, Lagier J-C, Parola P, Hoang VT, Meddeb L, Mailhe M, et al. Hydroxychloroquine and azithromycin as a treatment of COVID-19: results of an open-label non-randomized clinical trial. International Journal of Antimicrobial Agents. 2020 Mar;105949.

38. Gautret P, Lagier J-C, Parola P, Hoang VT, Meddeb L, Sevestre J, et al. Clinical and microbiological effect of a combination of hydroxychloroquine and azithromycin in 80 COVID-19 patients with at least a six-day follow up: A pilot observational study. Travel Medicine and Infectious Disease. 2020 Mar;34:101663.

39. Mahevas M, Tran V-T, Roumier M, Chabrol A, Paule R, Guillaud C, et al. No evidence of clinical efficacy of hydroxychloroquine in patients hospitalized for COVID-19 infection with oxygen requirement: results of a study using routinely collected data to emulate a target trial [Internet]. Infectious Diseases (except HIV/AIDS); 2020 Apr [cited 2020 May 28]. Available from: http://medrxiv.org/lookup/doi/10.1101/2020.04.10.20060699

40. Tang W, Cao Z, Han M, Wang Z, Chen J, Sun W, et al. Hydroxychloroquine in patients mainly with mild to moderate COVID-19: an open-label, randomized, controlled trial [Internet]. Public and Global Health; 2020 Apr [cited 2020 May 28]. Available from: http://medrxiv.org/lookup/doi/10.1101/2020.04.10.20060558

41. Magagnoli J, Narendran S, Pereira F, Cummings T, Hardin JW, Sutton SS, et al. Outcomes of hydroxychloroquine usage in United States veterans hospitalized with Covid-19 [Internet]. Infectious Diseases (except HIV/AIDS); 2020 Apr [cited 2020 May 28]. Available from: http://medrxiv.org/lookup/doi/10.1101/2020.04.16.20065920

42. Radonovich LJ, Simberkoff MS, Bessesen MT, Brown AC, Cummings DAT, Gaydos CA, et al. N95 Respirators vs Medical Masks for Preventing Influenza Among Health Care Personnel: A Randomized Clinical Trial. JAMA. 2019 Sep 3;322(9):824.

43. Leung NHL, Chu DKW, Shiu EYC, Chan K-H, McDevitt JJ, Hau BJP, et al. Respiratory virus shedding in exhaled breath and efficacy of face masks. Nat Med. 2020 May;26(5):676–80.

44. McMichael TM, Clark S, Pogosjans S, Kay M, Lewis J, Baer A, et al. COVID-19 in a Long-Term Care Facility — King County, Washington, February 27–March 9, 2020. MMWR Morb Mortal Wkly Rep. 2020 Mar 27;69(12):339–42.

45. Fang Y, Nie Y, Penny M. Transmission dynamics of the COVID-19 outbreak and effectiveness of government interventions: A data-driven analysis. J Med Virol. 2020 Jun;92(6):645–59.

46. Gudbjartsson DF, Helgason A, Jonsson H, Magnusson OT, Melsted P, Norddahl GL, et al. Spread of SARS-CoV-2 in the Icelandic Population. N Engl J Med. 2020 Apr 14;NEJMoa2006100.

47. Lavezzo E, Franchin E, Ciavarella C, Cuomo-Dannenburg G, Barzon L, Del Vecchio C, et al. Suppression of COVID-19 outbreak in the municipality of Vo, Italy [Internet]. Epidemiology; 2020 Apr [cited 2020 May 28]. Available from: http://medrxiv.org/lookup/doi/10.1101/2020.04.17.20053157

48. Sutton D, Fuchs K, D’Alton M, Goffman D. Universal Screening for SARS-CoV-2 in Women Admitted for Delivery. N Engl J Med. 2020 May 28;382(22):2163–4.

49. Baggett TP, Keyes H, Sporn N, Gaeta JM. COVID-19 outbreak at a large homeless shelter in Boston: Implications for universal testing [Internet]. Public and Global Health; 2020 Apr [cited 2020 May 28]. Available from: http://medrxiv.org/lookup/doi/10.1101/2020.04.12.20059618

50. Moriarty LF, Plucinski MM, Marston BJ, Kurbatova EV, Knust B, Murray EL, et al. Public Health Responses to COVID-19 Outbreaks on Cruise Ships — Worldwide, February–March 2020. MMWR Morb Mortal Wkly Rep. 2020 Mar 27;69(12):347–52.

51. Liu Y, Ning Z, Chen Y, Guo M, Liu Y, Gali NK, et al. Aerodynamic analysis of SARS-CoV-2 in two Wuhan hospitals. Nature [Internet]. 2020 Apr 27 [cited 2020 May 28]; Available from: http://www.nature.com/articles/s41586-020-2271-3

